# A-to-I RNA editing in kidney tissue from patients with nephrotic syndrome

**DOI:** 10.64898/2026.08.08.26359884

**Authors:** Victoria Mezger, Michelle T. McNulty, Dongwon Lee, Matthew G. Sampson

**Author notes:** Corresponding author. Address: Division of Nephrology, Boston Children’s Hospital, Enders 509, 320 Longwood Avenue, Boston, Massachusetts 02115, USA.

## Abstract

**INTRODUCTION:** RNA editing has been implicated in endogenous double-stranded RNA (dsRNA) sensing and inflammatory disease, but its prevalence, genetic regulation, and consequences in diseased human kidney tissue have not been systematically characterized. Because ADAR enzymes edit multiple neighboring adenosines often in the same transcript, analyzing these sites holistically (as “clusters”) may reveal effects missed by single-site analysis.

**METHODS:** We profiled both single-site and cluster A-to-I RNA editing in the kidneys of 215 participants from the Nephrotic Syndrome Study Network with focal segmental glomerulosclerosis or minimal change disease who had microdissected glomerular and/or tubulointerstitial RNA-seq and blood genome sequencing.

We tested single-site and cluster editing association with estimated glomerular filtration rate, proteinuria, and an interferon-stimulated gene expression score. To discover the genetic determinants of editing, we conducted mapping of both single-site, *cis*-editing QTL and cluster-level editing QTLs (cledQTLs). We then tested cledQTLs for colocalization with kidney eQTLs and kidney-relevant GWAS.

**RESULTS:** Greater cluster mean editing in tubulointerstitium was associated with lower interferon-stimulated gene activity (*P* = 4.81 × 10^-9^), less proteinuria (*P* = 0.01) and higher eGFR (*P* = 8.52 × 10^-5^). Genetic mapping identified 290 glomerular and 473 tubulointerstitial single-site edQTLs, as well as 21 glomerular and 51 tubulointerstitial cledQTLs. We identified 10 colocalized signals between cledQTL and GWAS and 14 between cledQTL and eQTL. Nine of 51 tubulointerstitial cledQTL clusters were individually associated with eGFR in NEPTUNE.

**CONCLUSION:** These results identify A-to-I RNA editing as a measurable and partly genetically regulated molecular phenotype in proteinuric kidney disease and nominate clustered editing of tubulointerstitial transcripts as a putative contributor to attenuated immune activity and higher kidney function. Cluster-level analysis identified additional genetically regulated editing patterns and colocalized signals not detected at individual sites, highlighting the added value of analyzing nearby editing sites as clusters.

## INTRODUCTION

Adenosine-to-inosine (A-to-I) RNA editing is a post-transcriptional process that converts adenosines to inosines, read as guanosines by the cellular machinery^1^. A-to-I editing is catalyzed by the adenosine deaminases acting on RNA (ADARs) and occurs predominantly in double-stranded RNA, especially Alu-derived inverted repeats^1,2^. A-to-I editing can influence splicing, transcript stability, microRNA targeting, and coding sequences^1,3,4^.

ADAR-mediated editing is involved in innate immune control via altering RNA processing. Many human editing sites occur in Alu-derived inverted repeats and other dsRNA-forming transcript regions, providing endogenous ADAR substrates^1^. Insufficiently edited endogenous dsRNA can be sensed as non-self by MDA5 and trigger type I interferon signaling^5,6^. ADAR1-mediated editing helps prevent this response, and loss of ADAR1 function causes interferon-driven disease in mice and humans^5–7^. Recent work further suggests that ADAR1-dependent immune protection is concentrated in a limited subset of cytosolic dsRNA substrates rather than all transcripts capable of forming dsRNA^8^.

Local genetic variation also shapes RNA editing. RNA editing quantitative trait loci (edQTLs), variants influencing site-specific editing, have been mapped across human tissues and linked to immune-mediated diseases^9,10^. However, the landscape and genetic regulation of RNA editing in diseased kidney tissue remain less characterized. Proteinuric kidney diseases like focal segmental glomerulosclerosis (FSGS) and minimal change disease (MCD) are glomerular diseases caused in part by immune dysregulation that can have progressive kidney functional decline due to tubulointerstitial injury^11^. It remains unknown how A-to-I editing varies across glomerular (GLOM) and tubulointerstitial (TUBE) compartments in these diseases, and whether genetically regulated editing is associated with immune activity or kidney traits.

Here, we characterize A-to-I RNA editing in microdissected kidney tissue from 215 individuals with FSGS or MCD in the Nephrotic Syndrome Study Network (NEPTUNE)^12^, using GLOM and/or TUBE RNA-seq and blood genome sequencing (WGS). Because dsRNA regions often contain multiple nearby edited sites, we hypothesized that measuring editing across these “clusters” would be a more biologically informed model than considering single-site edits alone. We therefore tested mean cluster editing for associations with kidney function and interferon-stimulated gene activity. We mapped cis-genetic effects on both individual sites and clusters of editing sites, defining edQTLs and cluster-level editing QTLs (hereafter cledQTLs). Finally, we evaluated individual cledQTLs for clinical association and colocalization with kidney-relevant GWAS and eQTLs.

## METHODS

### Nephrotic Syndrome Study Network (NEPTUNE)

NEPTUNE is a multicenter longitudinal study of individuals with proteinuric glomerular disease^12^. Participants provided written informed consent and undergo clinically indicated kidney biopsy, with an additional research core collected when available. We analyzed NEPTUNE participants with biopsy diagnoses of FSGS or MCD and available kidney RNA-seq from microdissected GLOM and/or TUBE compartments and blood genome sequencing.

### RNA editing quantification

We quantified A-to-I RNA editing from duplicate-marked, genome-aligned RNA-seq BAM files at REDIportal hg38 candidate sites^13^. Editing levels were quantified from strand-aware site annotations, using an mpileup-based workflow adapted from the GTEx edQTL pipeline^9^. Pileups were generated using samtools mpileup with a GRCh38 reference and REDIportal site intervals, using uniquely mapped reads and base-quality > 20. For each sample and site, coverage was defined as the number of reads supporting either the annotated reference or edited base, and editing level was calculated as edited reads divided by coverage.

Sample-site editing fractions were retained when coverage was at least 10 reads. Sites were retained if an editing fraction of ≥0.10 was observed in at least 5% of samples within the corresponding compartment. Sites overlapping blood germline variants were excluded (**Fig. 1b**).

**Figure 1.**
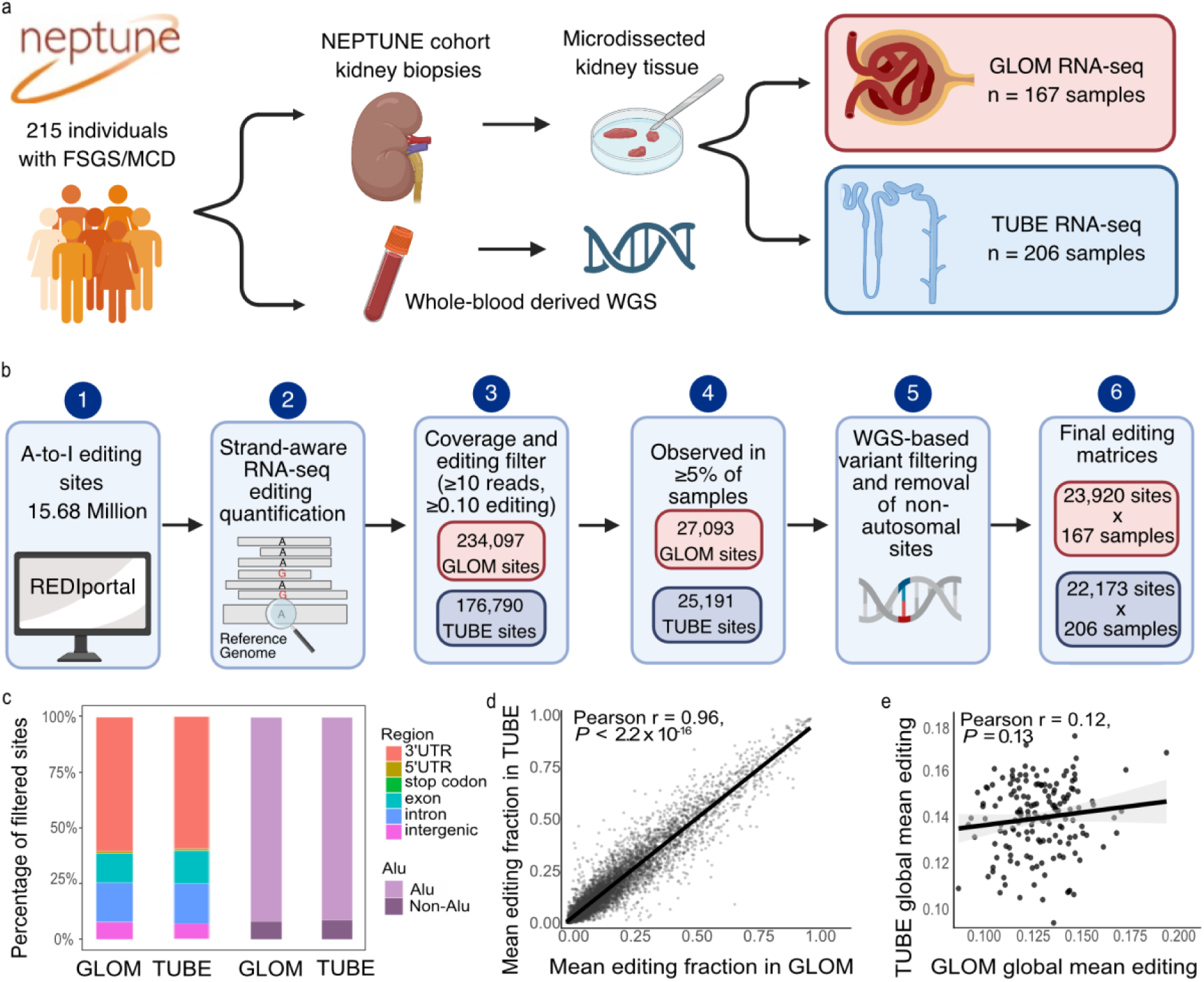
Study design and tissue-specific RNA editing landscape in NEPTUNE kidney. a. Overview of the NEPTUNE kidney RNA-seq dataset. Microdissected GLOM and TUBE RNA-seq samples from focal segmental glomerulosclerosis (FSGS) and minimal change disease (MCD) participants were analyzed together with whole-blood WGS genotypes. The dataset included 167 GLOM and 206 TUBE samples from 215 individuals, including 158 individuals with paired GLOM-TUBE data. Created in BioRender. Mezger, V. (2026) https://BioRender.com/rcm8u9i. b. Schematic workflow for strand-aware quantification and filtering of REDIportal hg38 A-to-I editing sites in GLOM and TUBE RNA-seq. Sites were filtered by read coverage, editing fraction, prevalence across samples and overlap with whole-blood WGS variants, yielding final matrices of 23,920 GLOM and 22,173 TUBE editing sites. Created in BioRender. Mezger, V. (2026) https://BioRender.com/rcm8u9i. c. Bars show the fraction of GLOM and TUBE editing sites assigned to genomic-region categories (left) and Alu status (right). Annotations are described in the **Supplementary Methods**. d. Concordance of mean editing across shared editing sites in GLOM vs TUBE. Each point represents one editing site present in both tissues, with mean editing calculated separately in GLOM and TUBE samples. The dashed diagonal indicates equal mean editing in GLOM and TUBE. Pearson correlation coefficient and two-sided *P*-value are shown. e. Concordance of sample-level global mean editing in GLOM vs TUBE. Each point represents one of 158 tissue-paired individuals. Line shows linear regression fit with 95% confidence interval. Labels report Pearson correlation coefficient and two-sided *P*-value.

Global mean editing was the mean across observed retained sites per sample and compartment.

### Mapping cluster-level editing, cledQTL, and edQTL

We constructed editing clusters from the single-site editing matrices separately in GLOM and TUBE. Distances were calculated between consecutive editing sites across the genome and clusters were defined as loci with at least 5 consecutive sites within 120nt from each other (**Fig. 2a)**.

**Figure 2.**
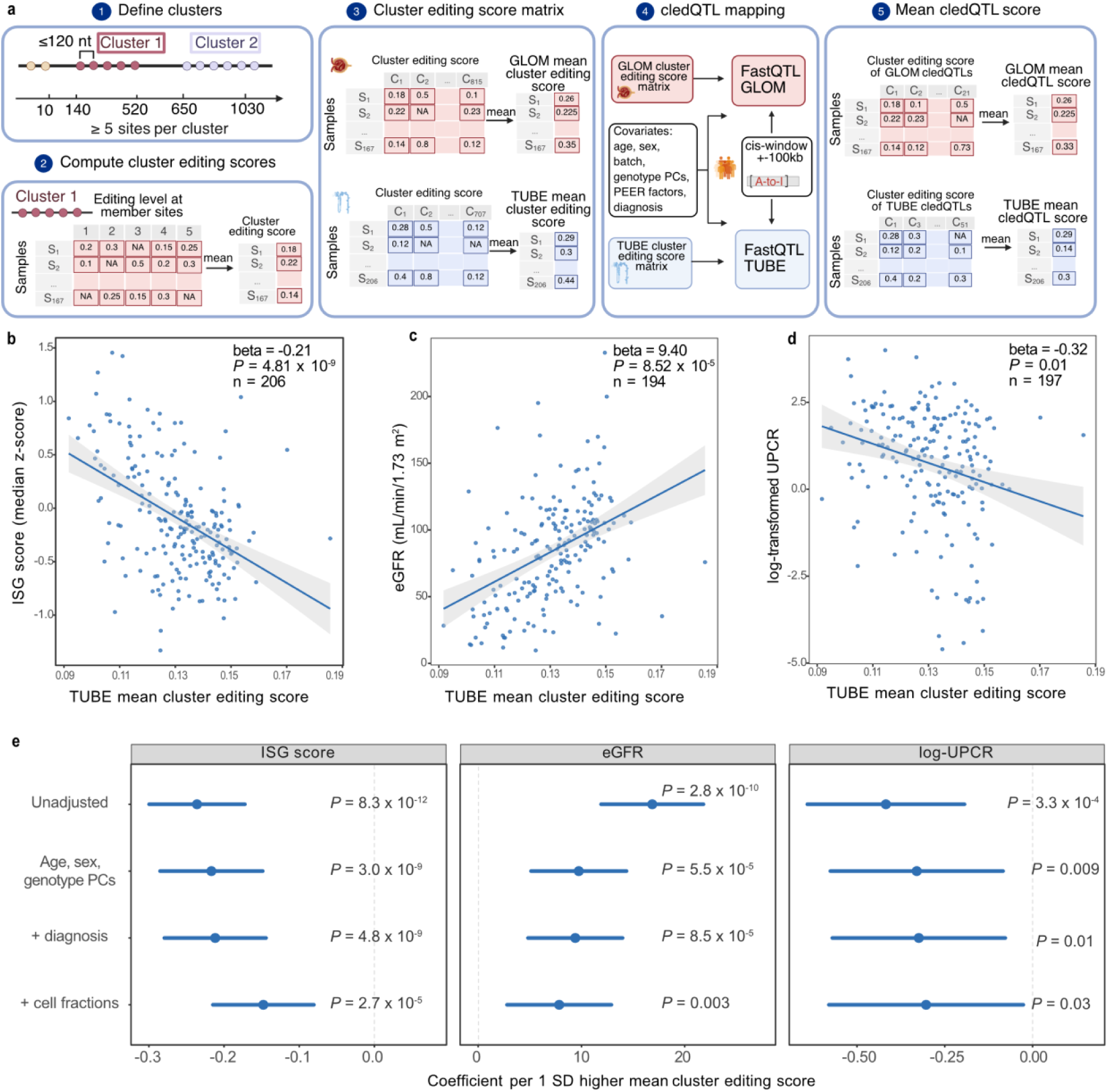
Clustered editing is associated with ISG activity and eGFR. a. Schematic of the cledQTL framework. Nearby edited sites were grouped into local editing-site clusters, summarized as per-sample mean cluster editing score, tested for *cis*-genetic regulation, and significant cledQTLs were used to calculate the mean cledQTL score. Created in BioRender. Mezger, V. (2026) https://BioRender.com/rcm8u9i. b. Association between TUBE mean cluster editing score and interferon-stimulated gene (ISG) activity. Each dot is a participant. The line shows the fitted linear regression with 95% confidence interval, adjusted for age, sex, genotype PCs, and histologic diagnosis. The coefficient is reported per 1-SD higher TUBE mean cluster editing score. c. Association between TUBE mean cluster editing score and eGFR at biopsy. The line shows the fitted linear regression with 95% confidence interval. The regression model adjusted for age, sex, genotype PCs, and histologic diagnosis. The displayed coefficient is reported per 1-SD higher TUBE mean cluster editing score. d. Association between TUBE mean cluster editing score and log-transformed UPCR. The line shows the fitted linear regression with 95% confidence interval. The regression model adjusted for age, sex, genotype PCs, and histologic diagnosis. The displayed coefficient is reported per 1-SD higher TUBE mean cluster editing score. e. Multivariable models of TUBE mean cluster editing score associations. Points show regression coefficients for the association of TUBE mean cluster editing score with ISG-score, UPCR and eGFR at biopsy; horizontal bars show 95% confidence intervals. Coefficients are reported per 1-SD higher TUBE mean cluster editing score. For ISG models, coefficients are in median ISG z-score units; for eGFR models, coefficients are in mL/min/1.73 m². Rows show unadjusted models and models sequentially adjusted for age, sex, genotype PCs, histologic diagnosis, and CIBERSORTx-derived TUBE cell-fraction estimates for proximal tubule, principal cell, thick ascending limb, and immune fractions. Nominal, two- sided *P*-values from the corresponding linear regression models are reported. The dashed vertical line indicates the null effect.

For each sample and cluster, the cluster editing score was defined as the mean editing fraction across observed member sites. At least two member sites with coverage of at least 10 reads were required to calculate a score; otherwise, the sample-cluster value was coded as missing.

For cledQTL mapping, clusters with ≤40% missingness across samples were retained. Remaining scores were mean-imputed and inverse-normal transformed.

We mapped cledQTLs using FastQTL^14^, adjusting for age, sex, genotype PCs 1-5, histologic diagnosis (FSGS vs MCD), RNA-seq batch (2 batches in GLOM, 3 in TUBE) and six PEER factors^15^. Variants with minor allele frequency (MAF) ≥0.05 were tested within ±100 kb of each cluster using 1000-10000 adaptive permutations. q-values were derived from beta-approximated permutation *P*-values using λ = 0.85, and significant cledQTL clusters were defined at q < 0.05. Single-site edQTLs were mapped analogously (**Supplementary Methods**). Significant cledQTLs were compared with single-site results and were considered to contain single-site edQTL evidence when at least one member site was significant at q < 0.05.

### Sample-level cluster editing scores, interferon-stimulated gene (ISG) scores, and clinical models

For each sample and compartment, the mean cluster editing score was defined as the mean of the cluster editing scores across all clusters tested, giving each cluster equal weight. The mean cledQTL score was defined similarly as the mean across all significant clusters from the cledQTL analysis.

We also compared mean editing between 11,646 sites within the 707 retained clusters with mean editing across the 10,527 remaining sites.

We summarized ISG activity using an external 331-gene ISG reference set^8^. Tissue-specific TPM expression matrices were transformed as log₂(TPM+1) and detected ISG genes were standardized across samples within each tissue. The per-sample ISG-score was defined as the median z-score across 313 detected ISGs.

Associations between (1) mean cluster editing score and (2) mean cledQTL score with (a) ISG-score, (b) urine protein creatinine ratio (UPCR), and (c) estimated glomerular filtration rate (eGFR) at biopsy were tested separately by tissue using linear regression adjusting for age, sex, genotype PCs 1-5, and diagnosis. Effects are reported per 1-SD higher z-standardized score.

To assess concentration of TUBE associations in cluster-forming regions, we fit pairwise joint linear regression models including both the mean cluster editing score and mean editing of sites not belonging to these clusters. TUBE sensitivity analyses additionally adjusted for inferred cell-type fraction using CIBERSORTx (**Supplementary Methods**).

### GWAS and eQTL colocalization

Significant edQTLs and cledQTLs were tested for colocalization with nine kidney/cardiometabolic GWAS (**Supplementary Table S6**) and matched-compartment NEPTUNE eQTLs using coloc.abf from the R package coloc with default settings, assuming at most one colocalized event per locus^16^.

Colocalization was performed using all overlapping variants within the cis-QTL window. Kidney eQTL analyses used GLOM and TUBE summary statistics from NEPTUNE^17^. Colocalization was defined as PP.H4 ≥ 0.8, indicating at least an 80% posterior probability that the two association signals share a causal variant within the tested regions. Details in **Supplementary Methods**.

## RESULTS

### A-to-I RNA editing landscape in microdissected proteinuric kidney tissue

We defined the landscape of A-to-I editing in 215 NEPTUNE participants with FSGS or MCD. There were 167 GLOM and 206 TUBE RNA-seq samples, including paired GLOM/TUBE data from 158 individuals (**Table 1**, **Fig. 1a**). We applied a strand-aware editing quantification workflow to the RNA-seq datasets (**Fig. 1b**), starting with a list of previously curated editing sites^13^. We used matched blood WGS to remove sites overlapping germline variants, reducing the risk that germline variation was misclassified as RNA editing. After excluding sites for coverage, editing level, and minimum prevalence across samples, and subsetting to autosomal sites, 23,920 GLOM and 22,173 TUBE editing sites were included in the final analytic set (**Supplementary Table S1)**. Genomic annotation showed the expected profile of ADAR-mediated A-to-I editing, with 60.33% of sites in GLOM and 59.32% in TUBE mapping to 3’UTRs, respectively and >90% overlapping Alu elements in both compartments (**Fig. 1c; Supplementary Table S2**). Although most sites mapped to noncoding regions, a small subset of RNA-editing events occurred in coding sequences with predicted protein-altering consequences. These events are described in the **Supplementary Results**.

**Table 1.** NEPTUNE analytic cohort and RNA editing dataset overview. Summary of RNA-seq sample counts, demographic characteristics, clinical variables, and diagnosis distribution for the NEPTUNE FSGS/MCD GLOM and TUBE analytic datasets. Continuous variables are reported as median (IQR), and categorical variables are reported as n (%), calculated among samples with available non-missing values. eGFR = estimated glomerular filtration rate; UPCR = urine protein to creatinine ratio; FSGS = focal segmental glomerulosclerosis; MCD = minimal change disease; EUR = European; AFR = African; AMR = Admixed American; EAS = East Asian; SAS = South Asian.

| Characteristic | Glomerular | Tubulointerstitial |
| --- | --- | --- |
| RNA-seq samples | 167 | 206 |
| **Demographics** |  |  |
| Age, median (IQR) | 19.0 (9.0-42.5); n=167 | 21.0 (11.0-46.8); n=206 |
| Female sex, n (%) | Female: 63/167 (37.7%) | Female: 82/206 (39.8%) |
| eGFR, median (IQR) | 89.3 (57.6-107.6); n=157 | 86.5 (55.5-106.2); n=194 |
| UPCR, median (IQR) | 2.7 (0.9-7.6); n=162 | 2.6 (0.9-6.8); n=200 |
| Pediatric age of kidney disease onset, n (%) | 77/146 (52.7%) | 94/180 (52.2%) |
| Adult age of kidney disease onset, n (%) | 69/146 (47.3%) | 86/180 (47.8%) |
| Age of kidney disease onset, median (IQR) | 16.5 (6.0-38.8); n=146 | 17.0 (6.0-39.5); n=180 |
| **Diagnosis** |  |  |
| FSGS, n (%) | 85/167 (50.9%) | 106/206 (51.5%) |
| MCD, n (%) | 82/167 (49.1%) | 100/206 (48.5%) |
| **Genotype inferred continental ancestry** |  |  |
| AFR, n (%) | 52/167 (31.1%) | 65/206 (31.6%) |
| AMR, n (%) | 33/167 (19.8%) | 41/206 (19.9%) |
| EAS, n (%) | 10/167 (6.0%) | 12/206 (5.8%) |
| EUR, n (%) | 59/167 (35.3%) | 70/206 (34.0%) |
| SAS, n (%) | 8/167 (4.8%) | 12/206 (5.8%) |
| UNKNOWN, n (%) | 5/167 (3.0%) | 6/206 (2.9%) |

Per sample, a median of 4,973 GLOM sites and 5,119 TUBE sites exceeded the ≥0.10 editing threshold, corresponding to 39.37% and 44.03% of observed sites, respectively. Across the 12,976 shared sites, mean editing levels were highly correlated between compartments (Pearson r = 0.96, *P <* 2.2 × 10^-16^; **Fig. 1d**). Per-sample global mean editing was only weakly correlated between compartments, not reaching statistical significance among paired samples (Pearson r = 0.12, *P* = 0.13; **Fig. 1e**) and varied with age in opposite directions across compartments, increasing with age in GLOM (rho = 0.45, *P* = 8.54 × 10^-10^) and decreasing with age in TUBE (rho = -0.31, *P* = 4.31 × 10^-6^; **Supplementary Fig. S1a**). No significant differences were observed by sex or inferred ancestry (**Supplementary Fig. S1b,c**).

We next examined associations with per-sample global mean editing. In covariate-adjusted models, a 1- SD higher global mean editing in TUBE was associated with a 7.90 mL/min/1.73 m² higher eGFR at biopsy (*P* = 1.10 × 10^-3^; **Supplementary Fig. S1d**). Reduced editing of endogenous dsRNA can also trigger MDA5-mediated interferon responses^5,8^. Consistently, global mean editing was also associated with a lower ISG-score in TUBE (β = -0.20 per 1-SD higher global mean editing, *P* = 1.01 × 10^-7^) (**Supplementary Fig. S1e**). Associations with UPCR were directionally consistent: a 1-SD higher global mean editing fraction in TUBE was associated with lower UPCR at biopsy (β = -0.26, *P* = 0.046) (**Supplementary Fig. S1f**). The corresponding GLOM associations were not significant.

Together, these analyses establish A-to-I RNA editing as a measurable, clinically associated phenotype in microdissected nephrotic syndrome kidney tissue and motivated subsequent analyses of coordinated editing across local clusters, individual editing sites, and genetic regulation of both.

### Clustered tubulointerstitial editing is associated with lower ISG activity, higher eGFR, and lower proteinuria

Because endogenous dsRNA sensing depends on RNA duplex structure^5,18^, the overall edited state of a dsRNA region may be more relevant to MDA5-mediated immune activation than editing at any single nucleotide alone^8^. This motivated us to also analyze nearby editing sites as a coordinated group of editing events, or “clusters”, to capture the biological phenomenon of linked editing and potential consequences.

We defined primary clusters by grouping adjacent filtered editing sites within 120 nucleotides (nt), requiring at least five sites per cluster as in a previous analysis^8^. Clusters contained a median of 11 sites and spanned approximately 180 bp. After requiring no more than 40% missing sample-level cluster values, 815 GLOM and 707 TUBE clusters remained (**Fig. 2a**). Among these, 442 cluster overlaps were observed between GLOM and TUBE, defined by at least one shared editing site.

Higher global mean editing in GLOM was not associated with lower ISG activity. We therefore calculated a mean cluster editing score only across all 707 common TUBE clusters and tested for association with ISG activity. In TUBE, higher mean cluster editing score was associated with lower ISG activity after adjusting for age, sex, genotype PCs and diagnosis (β = -0.21 ISG-score units per 1-SD higher mean cluster editing score; *P* = 4.81 × 10⁻^9^; **Fig. 2b**). Higher TUBE mean cluster editing score was also significantly associated with participants’ kidney function. In a covariate-adjusted model, higher TUBE mean cluster editing score was associated with higher eGFR (β = 9.40 mL/min/1.73 m^2^ per 1-SD higher TUBE mean cluster editing score; *P* = 8.52 × 10⁻^5^; **Fig. 2c**) and lower UPCR (β = -0.32; *P* = 0.01, **Fig. 2d**). Because tubulointerstitial bulk RNA-seq reflects variation in cell composition, we additionally adjusted for inferred proximal tubule, principal cell, thick ascending limb, and immune-cell fractions using CIBERSORTx^19^. All associations remained significant (ISG: β = -0.15, *P* = 2.65 × 10^-5^; eGFR: β = 7.85, *P* = 0.003; UPCR: β = -0.30; *P* = 0.03; **Fig. 2e**).

To determine whether these associations were stronger when comparing clusters versus unclustered editing sites, we compared the mean cluster editing score with the mean editing across the 10,527 autosomal sites that did not belong to these filtered clusters, defined as “outside-cluster mean editing” (**Methods**). In models including both scores, the mean cluster editing score remained associated with lower ISG activity (β = -0.25; *P* = 2.36 × 10⁻^8^) and higher eGFR (β = 11.68; *P* = 8.15 × 10⁻^5^), whereas the outside-cluster mean editing was not independently associated with either outcome (ISG: β = 0.06, *P* = 0.14; eGFR: β = -3.77, *P* = 0.19). The same pattern was observed for UPCR: cluster editing remained associated with lower UPCR (β = -0.39; *P* = 0.02), whereas outside-cluster editing was not (β = 0.10; *P* = 0.51). These findings localize the TUBE editing signal associated with lower ISG activity, proteinuria and higher eGFR to sites within local editing clusters.

### *Cis*-edQTL and -cledQTL mapping

*Cis*-genetic effects on A-to-I editing have been described across human tissues^9,10^, yet their role in kidney has not been systematically characterized. To address this, we mapped genetic effects on single-site RNA editing levels using FastQTL^14^. Per-site editing levels were filtered for missingness and variance, mean-imputed, inverse-normal transformed, and tested against germline variants within ±100 kb around each edited site, accounting for covariates and six PEER factors^15^. After applying a missingness filter of at most 40% samples missing per site^9^, 9,888 GLOM (41.3% of 23,920 considered earlier) and 8,961 TUBE (40.4% of 22,173) autosomal sites entered edQTL mapping.

We define edQTL sites as editing sites with at least one significant *cis* association and edGenes as genes annotated to edQTL sites. At q < 0.05, we identified 290 edQTL sites in GLOM and 473 in TUBE, corresponding to 2.9% and 5.3% of tested sites, respectively. As multiple edQTLs mapped to the same gene, the edQTLs overall mapped to 141 GLOM and 180 TUBE edGenes. Lead variants for edQTLs were closer to their corresponding editing sites than expected under a per-site matched null (mean distance GLOM: 13.8 kb observed vs 48.9 kb expected; TUBE: 18.1 kb observed vs 48.6 kb expected; empirical *P* < 1 × 10^-4^ for both; **Fig. 3a**). 93.6% of cross-compartment effects had the same direction (*P* = 5.74 × 10^-73^, **Fig. 3b**). These results show that a subset of single-site RNA editing is locally genetically regulated in nephrotic syndrome kidney tissue, with many effects shared between GLOM and TUBE.

**Figure 3.**
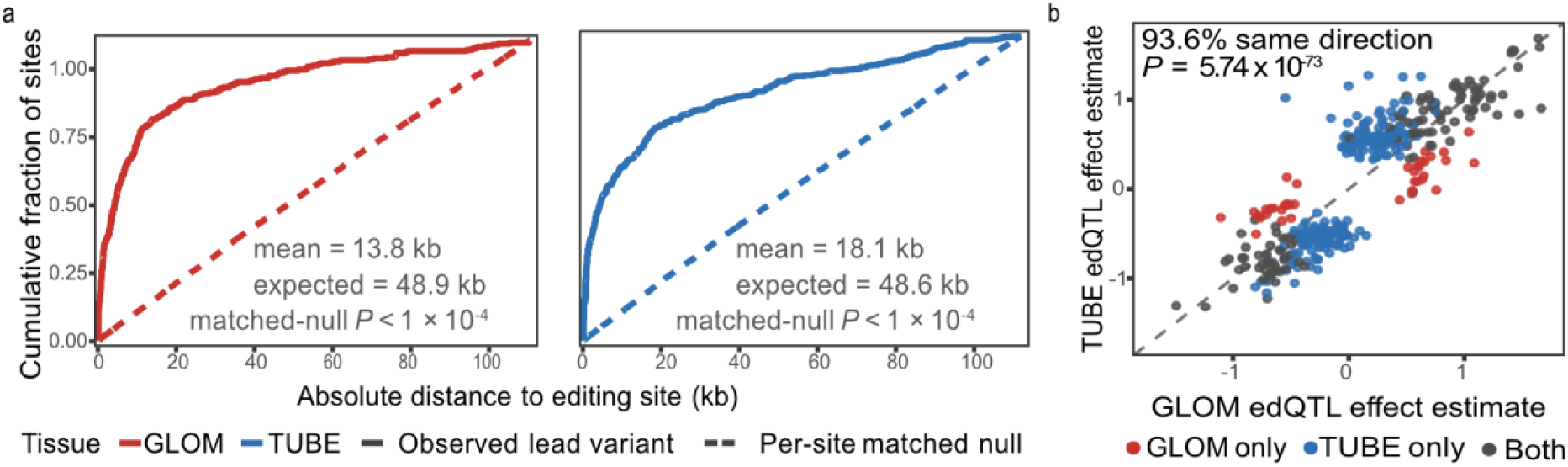
Kidney RNA editing is genetically regulated in *cis*. a. Distance between each significant single-site edQTL and its lead variant, defined as the *cis*-variant with the smallest FastQTL *P*-value for that editing site. Solid lines show the cumulative distribution of absolute distances between lead variants and editing sites. Dashed lines show the cumulative distribution expected under a per-site matched null procedure in which control variants were sampled from the same tested *cis* window. Text reports the observed, the average mean distance across 10,000 matched-null resamples, and the two-sided empirical *P*-value and the two-sided empirical *P*-value comparing the observed mean distance with the matched-null distribution. Red and blue represent GLOM and TUBE, respectively. b. Cross-compartment concordance of single-site edQTL effects. Each point represents one variant-RNA editing site pair that was tested in both GLOM and TUBE and was significant at q < 0.05 in at least one compartment, with color indicating the significant compartment. 369 pairs are shown, with 39 significant in GLOM only, 207 in TUBE and 123 significant in both. The axes show the estimated change in normalized editing level per additional copy of the tested allele in GLOM and TUBE, respectively. Positive values indicate higher editing and negative values indicate lower editing with increasing allele dosage. The dashed diagonal indicates equal effect estimates, and gray lines indicate no effect.

Prior studies have shown that local genetic variants can affect multiple editing sites within the same region^9^. Consistent with this, 179 of 290 GLOM edQTL sites (61.7%) and 335 of 473 TUBE edQTL sites (70.8%) were located within 120 nt of another edQTL site. This motivated us to test for genetic effects associated with the mean editing level of each cluster in the filtered datasets, which we refer to as cledQTLs. At q < 0.05, we identified 21 significant GLOM and 51 significant TUBE cledQTLs, respectively, with seven overlapping between compartments (**Supplementary Table S3**). The majority (20/21 and 38/51, respectively) of the cledQTLs contained at least one significant single-site edQTL member, indicating that cluster-level discovery captures and extends the single-site signal.

### Associations of genetically regulated editing sites and clusters

Similar to associations seen amongst all clusters (including those without a cledQTL), a higher mean editing score restricted to the 51 significant TUBE cledQTLs was associated with lower ISG activity (β = -0.20, *P* = 1.16 × 10^−8^) and higher eGFR (β = 8.16, *P* = 6.18 × 10^−4^) (**Supplementary Fig. S2**). These findings place the cledQTLs within the broader TUBE clustered-editing state while providing direct genetic support for locus-level follow-up. We therefore examined individual cledQTL clusters for clinical associations. Beyond this, with knowledge of the variant comprising the editing QTLs, we could test for colocalization with GWAS and eQTLs.

In a model testing each of the 72 significant cledQTLs for association with eGFR or UPCR, adjusting all models for age, sex, diagnosis and genotype PCs, 9 of 51 TUBE clusters were significantly associated with eGFR (**Fig. 4a**; **Supplementary Table S4**). No GLOM cluster was significant.

**Figure 4.**
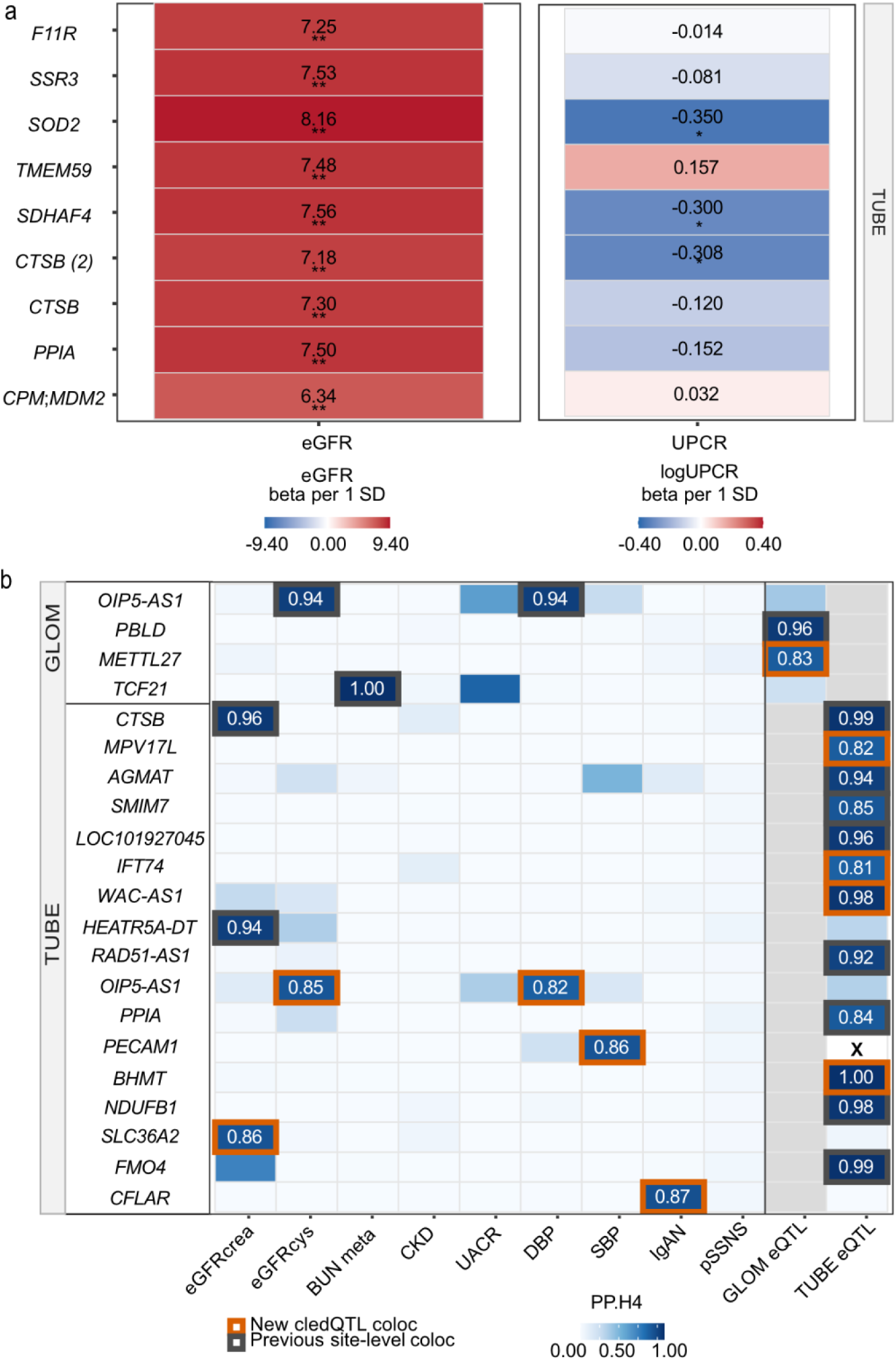
Clinical associations and colocalization of kidney cledQTLs. a. Associations of observed cluster editing score with eGFR at biopsy and UPCR in NEPTUNE, estimated separately for significant TUBE cledQTL clusters. Rows represent genes assigned to individual cledQTL clusters. Values inside cells indicate the estimated outcome difference per 1-SD higher cluster editing score from linear regression models adjusted for age, sex, histologic diagnosis, and genotype PCs. For eGFR, coefficients are reported in mL/min/1.73 m²; for UPCR, coefficients are reported on the natural-log scale. Cell fill indicates signed effect size. Asterisks denote nominal significance (\**P* < 0.05) and Benjamini-Hochberg significance (**FDR < 0.05) within each compartment-outcome analysis. b. Colocalization of significant kidney cledQTLs with GWAS traits and matched-compartment kidney eQTLs. Rows represent significant cledQTL clusters with at least one GWAS or eQTL colocalization at PP.H4 ≥ 0.8. Columns show kidney-relevant GWAS traits and matched-compartment GLOM or TUBE eQTLs. Cell shading indicates PP.H4, the posterior probability that the cledQTL and the corresponding GWAS or eQTL association share a causal signal; numeric values are shown only for cells with PP.H4 ≥ 0.8. Orange outlines denote colocalizations newly identified at the cledQTL level, whereas black outlines denote colocalizations also observed in the corresponding single-site edQTL analysis. White cells with X indicate matched-compartment eQTL colocalizations that could not be evaluated. Grey cells indicate the non-matched tissue compartment, which was not tested. GWAS trait abbreviations: eGFRcrea, creatinine- based estimated glomerular filtration rate; eGFRcys, cystatin C-based estimated glomerular filtration rate; BUN, blood urea nitrogen; CKD, chronic kidney disease; UACR, urinary albumin-to-creatinine ratio; DBP, diastolic blood pressure; SBP, systolic blood pressure; IgAN, IgA nephropathy; pSSNS, pediatric steroid-sensitive nephrotic syndrome.

For comparison, we applied the same clinical-association framework to significant single-site edQTLs. In TUBE, one edQTL site was associated with eGFR and five with UPCR (**Supplementary Table S5**). Notably, cluster-level analysis identified additional eGFR associations without corresponding significant single-site edQTL associations, including clusters annotated to Transmembrane Protein 59 (*TMEM59)*, an autophagy regulator, and Signal Sequence Receptor Subunit 3 (*SSR3),* a subunit of the ER translocon-associated protein complex **(Supplementary Table S4)**^20,21^. This suggests aggregation across coordinated sites can reveal associations not apparent from individual sites alone.

We then tested whether cledQTLs colocalized with kidney-relevant GWAS. We identified 10 cledQTL- GWAS colocalization events across 8 unique clusters using a PP.H4 threshold of 0.8 (**Fig. 4b; Supplementary Table S4**). Compared with the single-site edQTL-GWAS colocalization results (**Supplementary Table S5**), the cledQTL analysis nominated additional clusters annotated to OIP5 Antisense RNA 1 *(OIP5-AS1),* Solute Carrier Family 36 Member 2 *(SLC36A2),* Platelet and Endothelial Cell Adhesion Molecule 1 *(PECAM1),* and CASP8 and FADD-like Apoptosis Regulator *(CFLAR)*. Again, cledQTL colocalization identified significant signals that would not have been found by single-site edQTL-GWAS analysis.

We also tested colocalization of clusters and GLOM/TUBE eQTLs and identified 14 significant signals (**Fig. 4b**). Compared with single-site edQTL-eQTL results (**Supplementary Table S5**), the cledQTL analysis nominated several additional eQTL-linked loci, including WAC Antisense RNA 1 (*WAC-AS1*) and Mitochondrial Inner Membrane Protein Like V17 (*MPV17L*) in TUBE. Both have been previously implicated in renal tubular stress and injury pathways^22,23^. Among the cledQTL colocalization candidates, Cathepsin B (*CTSB*) cluster 2, the second of two significant TUBE *CTSB* clusters, was the only cledQTL that colocalized with both a GWAS (creatinine-based eGFR) and an eQTL, and was also associated with higher eGFR (**Fig. 5**). The cledQTL lead variant was associated with both the cluster editing and several member sites, providing convergent evidence linking local genetic regulation of RNA editing, *CTSB* expression, and kidney function. Together with the established role of *CTSB*, a lysosomal cysteine protease implicated in kidney injury pathways involving autophagy, apoptosis, and inflammation^24,25^, this convergence prioritizes *CTSB* for causal and mechanistic follow-up.

**Figure 5.**
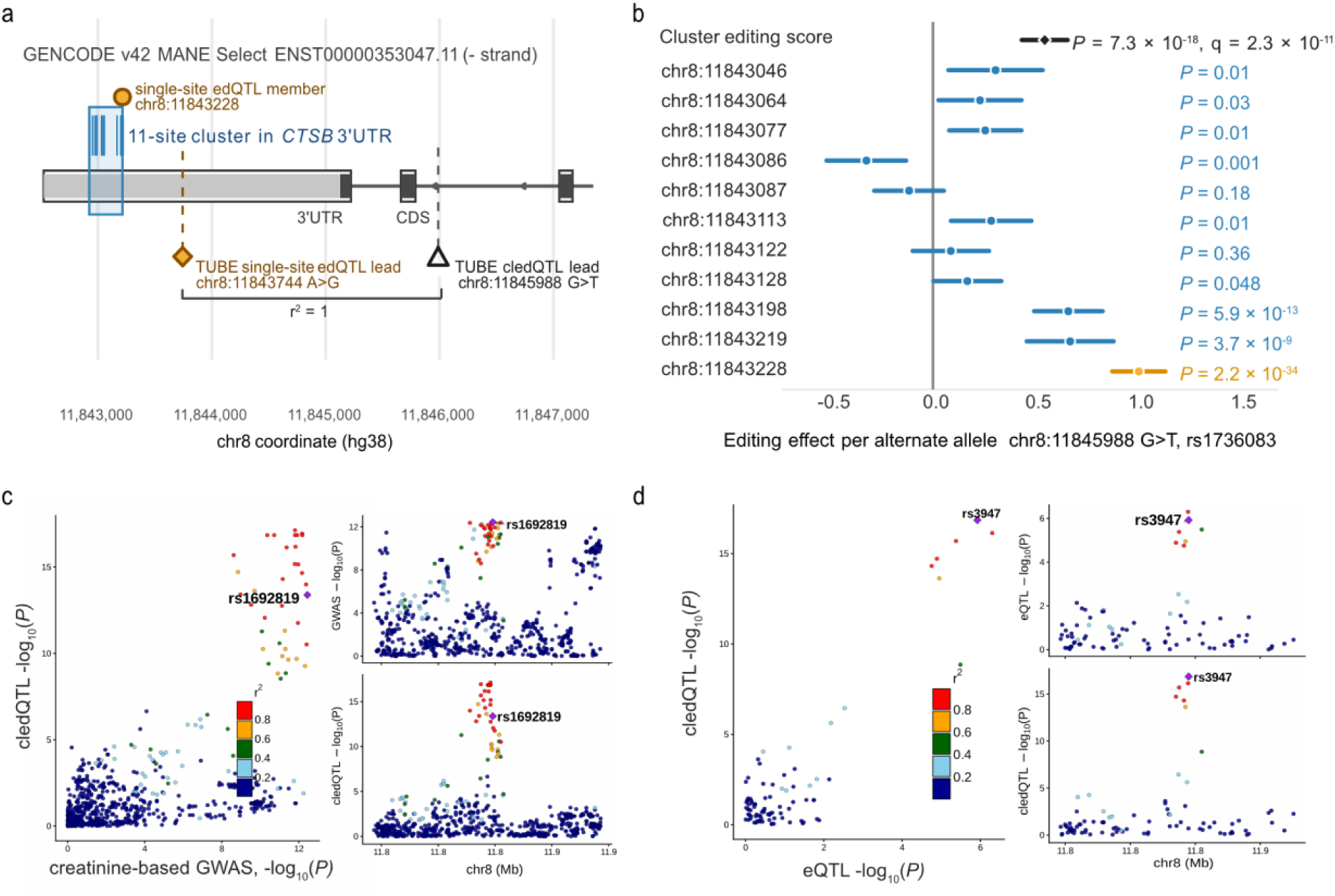
Clinical and genetic evidence at the TUBE *CTSB* cledQTL locus. a. *CTSB* transcript structure and location of the TUBE *CTSB* cledQTL cluster. Local map of the GENCODE v42 canonical/MANE Select *CTSB* transcript ENST00000353047.11. Gray boxes indicate UTRs, dark boxes indicate the coding sequence, black-outlined boxes indicate exons, and arrows indicate transcript orientation. The blue shaded region marks the 11-site TUBE *CTSB* cledQTL cluster, which spans chr8:11843046-11843228 and lies within the *CTSB* 3’UTR; blue ticks indicate individual member editing sites. The gold circle marks the cluster-member editing site chr8:11843228 that was also significant in the single-site edQTL analysis. The gold diamond marks the single-site edQTL lead variant and the white triangle marks the TUBE cledQTL lead variant. The two variants were in perfect linkage disequilibrium (r^2^ = 1) in the NEPTUNE TUBE genotype panel and therefore represent the same underlying association signal. b. Effects of the cledQTL lead variant on cluster mean editing and individual cluster-member sites. Effects are coded per alternate T allele. The black diamond shows the cluster-level effect on mean editing across the 11-site *CTSB* cluster. Blue points show effects on individual cluster-member editing sites. The orange point marks the cluster-member editing site that was also significant in the single-site edQTL analysis. Horizontal bars show 95% confidence intervals. The vertical line indicates no effect. The cluster-level q value is the FastQTL FDR-adjusted q value; member-site labels show nominal *P*-values for the same variant-site association. c. Scatter and regional association plots for the TUBE *CTSB* cledQTL and creatinine-based eGFR GWAS. The left panel shows -log_10_(*P*) values for the cledQTL association plotted against -log_10_(*P)* values for the eGFR GWAS association across variants in the locus. Right panels show regional association signals for the eGFR GWAS and the TUBE *CTSB* cledQTL. Points are colored by linkage disequilibrium, r², with the rs1692819 variant. LD was calculated from the genotypes of NEPTUNE participants. Only variants available and harmonized in both datasets are shown. d. Scatter and regional association plots for the TUBE *CTSB* cledQTL and matched-compartment *CTSB* eQTL. The left panel shows -log_10_(*P*) values for the cledQTL association plotted against -log_10_(*P*) values for the *CTSB* eQTL association across variants in the locus. Right panels show regional association signals for the *CTSB* eQTL and the TUBE *CTSB* cledQTL. Points are colored by linkage disequilibrium, r², with the rs3947 variant. LD was calculated from the genotypes of NEPTUNE participants. Only variants available and harmonized in both datasets are shown.

## DISCUSSION

To test the hypothesis that A-to-I RNA editing, an established post-transcriptional regulatory mechanism with a known role in attenuating immune responses, contributes to the pathogenesis and/or progression of proteinuric kidney diseases, we mapped the landscape of glomerular and tubulointerstitial editing in microdissected kidney tissue from participants with FSGS or MCD and related it to clinical outcomes and molecular endophenotypes. Our most striking findings were that (1) as opposed to analysis of editing sites individually, grouping them into clusters of neighboring sites arguably provides a more biologically accurate way to model editing and that (2) increased editing in the tubulointerstitium (but not the glomerulus) was associated with higher eGFR, less proteinuria, and lower intrarenal expression of interferon signaling genes.

Our use of a clustering approach was motivated by a 2025 study by Sun et al., who demonstrated in a 293T cell line that quantifying editing as clusters (as opposed to by single sites) provided a more accurate model of the immunogenicity of dsRNA^8^. Here, we validate this metric in human kidney tubulointerstitium, finding, in joint models with non-overlapping editing-site sets that the cluster editing score remained associated with eGFR, UPCR and ISG, whereas editing outside of clusters did not. These findings suggest that the associations between higher editing, lower immune activity and lower kidney disease severity are concentrated within clustered editing regions rather than reflecting editing across the transcriptome more broadly. This is consistent with prior work showing that MDA5-dependent immunogenicity is carried by a limited subset of cytosolic dsRNAs^8^. Although our coordinate-defined clusters are not validated dsRNA structures, they may enrich for RNA regions in which clustered A-to-I editing influences innate immune sensing and kidney injury.

From the perspective of analysis of human data, our work does not suggest that considering individual site editing should be discarded. However, it does suggest that future editing-phenotype association studies may be less accurate if a metric of cluster editing is not included. From an experimental perspective, it would be seemingly important to evaluate differences in cellular behavior arising from introducing an edit to an individual site versus a complete cluster.

It is notable that significant associations between editing and molecular and clinical phenotypes were only observed in TUBE. Several explanations are possible. From a technical perspective, the cell types making up the TUBE samples may better capture epithelial injury and immune-response states that are visible in bulk RNA-seq, whereas glomerular effects may be restricted to specific cell types, transcripts, or injury states not readily observed in microdissected bulk tissue. From a biologic perspective, given that tubulointerstitial injury and immune-response programs are key components of the kidney damage from proteinuric kidney diseases, it is plausible that the tubulointerstitial compartment is the predominant site where RNA editing and interferon biology intersect in these diseases^26^. However, GLOM editing should not be dismissed as biologically irrelevant. Prior work has shown ADAR-mediated editing of APOL1 dsRNA in podocytes and glomerular tissue, supporting glomerular relevance for specific transcripts^27^. Finally, from a statistical perspective, TUBE also had a larger sample size which may have increased power and discovery of more significant single-site and cluster-level edQTLs. Future studies in larger sample sizes and those that have single cell/nuclear RNA-seq data will help us to better understand the extent of glomerular editing’s role in disease.

Finally, our GLOM and TUBE editing QTL analysis identified 72 cledQTLs and more than 700 edQTLs. Here, we used these discoveries for both cluster-level and single-site colocalization, pairing them with varying degrees of orthogonal support to provide complementary evidence linking genetically regulated RNA editing to kidney disease. *CTSB* arguably represented the locus with strongest convergent support. It was a TUBE cledQTL, where increased editing was associated with higher eGFR. Furthermore, it colocalized with a GWAS of eGFR and a TUBE eQTL. This raises the testable hypothesis that a genetic variant alters editing levels of the *CTSB* locus in a manner that changes the gene’s expression and alters the eGFR.

At the cluster level, additional edQTLs such as *SLC36A2* and *CFLAR* suggest that modeling editing as a coordinated set of editing sites within a restricted region may more accurately capture genomic biology not prioritized by single-site edQTL mapping alone. *CFLAR* encodes c-FLIP, a regulator of death-receptor and caspase-8-mediated apoptosis that is expressed in kidney cortex and tubular epithelial cells and has been implicated in cytokine-induced tubular epithelial injury^28,29^. *SLC36A2* encodes proton-coupled amino acid transporter 2 (*PAT2*), a transporter involved in renal handling of glycine, proline, and hydroxyproline. Human genetic studies of iminoglycinuria and hyperglycinuria have implicated *SLC36A2*/*PAT2* in renal tubular reabsorption of these amino acids, and *SLC36A2* has been described at the apical surface of the human renal proximal tubule^30^. Together, these results argue that RNA editing can add a post-transcriptional layer to kidney genetic interpretation.

Several limitations remain. First, bulk RNA-seq from microdissected tissue cannot identify the precise cell types or injury states driving the TUBE cluster-editing signal. Single-cell, single-nucleus, and/or spatial kidney datasets will be needed to determine whether these signals arise from proximal tubule cells, other epithelial cells, immune-cells, or coordinated epithelial-immune interactions. Second, although the cledQTL framework is motivated by dsRNA biology, the clusters are coordinate-defined editing site groups and not experimentally validated RNA structures. Direct validation of dsRNA formation, ADAR dependence, and MDA5 activation would strengthen our mechanistic understanding. Furthermore, the detected cledQTLs likely represent only a subset of genetically regulated clusters, because limited sample size, variable RNA-seq coverage, and multiple-testing burden reduce power to detect weaker effects.

Larger kidney datasets may therefore identify additional genetically regulated clusters. Third, colocalization identifies shared local genetic signals but does not establish mediation or causal direction. Finally, replication in an independent nephrotic syndrome cohort with matched RNA-seq, genotype data, and clinical follow-up remains important. However, it is difficult to find independent cohorts that have a similar collection of blood and tissue-genomics profiles and clinical data.

In conclusion, these findings support A-to-I RNA editing as a partly genetically regulated post-transcriptional phenotype in proteinuric kidney disease. They nominate increased TUBE cluster editing as protective factor in these diseases. Finally, the edQTL and cledQTL regulatory variants can now be examined in independent human cohorts and biobanks, or taken to the bench, to establish causal inferences both epidemiologically and biologically.

## Data Availability

Raw WGS and RNA-seq data from GLOM and TUBE are available through NEPTUNE (https://www.neptune-study.org/ancillary-studies). Single cell RNA-seq data used to infer cell-type proportions can be accessed from KPMP repository (https://atlas.kpmp.org/repository). eQTL data are available through NephQTL2 (https://www.nephqtl2.org).

## Acknowledgments

We would like to thank Jin Billy Li and Qin Li for the insightful discussions during the development of this study. We would like to thank all members from the Sampson and Lee labs for their helpful comments on this study. We would like to acknowledge the Boston Children’s Hospital High-Performance Computing Resources BCH HPC Clusters, Enkefalos 3 (E3), made available for conducting the research reported in this publication.

The Nephrotic Syndrome Study Network (NEPTUNE) is alumni of the Rare Diseases Clinical Research Network (RDCRN), which is funded by the National Institutes of Health (NIH) and led by the National Center for Advancing Translational Sciences (NCATS) through its Division of Rare Diseases Research Innovation (DRDRI). NEPTUNE has been funded under grant number U54DK083912 as a collaboration between NCATS and the National Institute of Diabetes and Digestive and Kidney Diseases (NIDDK). Additional funding and/or programmatic support is provided by the University of Michigan, NephCure Kidney International, Alport Syndrome Foundation, and the Halpin Foundation. RDCRN active consortia and alumni are supported by the RDCRN Data Management and Coordinating Center (DMCC), funded by NCATS and the National Institute of Neurological Disorders and Stroke (NINDS) under U2CTR002818.

The Kidney Precision Medicine Project (KPMP) is supported by the National Institute of Diabetes and Digestive and Kidney Diseases (NIDDK) through the following grants: U01DK133081, U01DK133091, U01DK133092, U01DK133093, U01DK133095, U01DK133097, U01DK114866, U01DK114908, U01DK133090, U01DK133113, U01DK133766, U01DK133768, U01DK114907, U01DK114920, U01DK114923, U01DK114933, U24DK114886, UH3DK114926, UH3DK114861, UH3DK114915, and UH3DK114937. We gratefully acknowledge the essential contributions of our patient participants and the support of the American public through their tax dollars.

## Author Contributions

V.M. and M.T.M designed and performed statistical analyses and wrote the manuscript.

D.L. designed statistical analyses and supervised the overall study.

M.G.S. conceived and supervised the overall study and wrote and edited the manuscript.

All authors discussed results and participated in editing and preparation of the manuscript

