## Supplement for "A-to-I RNA editing in kidney tissue from patients with nephrotic syndrome"

**This file includes:**

Supplementary Methods

Supplementary Results

Figures S1 to S2

Tables S1 to S7

Supplementary Text

Supplementary References

**SUPPLEMENTARY METHODS**

### **Processing of NEPTUNE Whole Genome Sequencing**

Whole-genome sequencing was performed on whole-blood DNA from 864 NEPTUNE participants. Libraries were sequenced on the Illumina HiSeq 4000 platform to a mean genome-wide coverage of approximately 30X. FASTQ files were assessed for read quality using FastQC v0.11.9^S1^ and FastQ Screen v0.15.1^S2^, and adapter and quality trimming was performed using fastp v0.24.0^S3^. Reads were aligned to the GRCh38 (hg38) human reference genome using BWA-MEM v0.7.17^S4^. Joint variant calling and filtering were performed using GATK v4.1.8.0^S5^ following the GATK germline short-variant discovery best-practices workflow.

For this study, NEPTUNE hg38 whole-blood WGS VCFs were processed separately for the GLOM and TUBE analytic sample sets. Source records were restricted to variants with InbreedingCoeff > -0.3, multiallelic records were split, genotypes with genotype quality (GQ) < 20 were set to missing, and no samples were removed using the >10% genotype-missingness threshold. Variants were removed if they had minor allele frequency < 0.01, genotype call rate < 85%, or Hardy-Weinberg equilibrium

*P* < 1 × 10^-6^. The final genotype VCFs contained 14,532,906 (GLOM) and 13,885,544 (TUBE) variants.

Genetic ancestry was inferred from WGS data using peddy^S6^. For population adjustment, genotype principal components were computed separately for the GLOM and TUBE sample sets using PLINK. Variants were LD-pruned using a 50-variant window, a 10-variant step and removal of variants with r^2^ > 0.1. The first five principal components were used as covariates in downstream PEER, cledQTL, edQTL and regression analyses.

**Processing of NEPTUNE bulk RNA-seq**

We used previously generated NEPTUNE microdissected kidney RNA-seq alignment files and expression matrices from the NEPTUNE ASE pipeline^S7^. Briefly, QC was performed using FastQC. Reads were aligned using STAR v.2.7.11b and duplicate reads were marked using samtools markdup v1.19.2. Gene quantification was performed using RSEM v1.3.1 with the hg38 reference genome, GENCODE v45.

### **Region annotation**

RNA editing sites were annotated using hg38 REDIportal-derived site annotations and repeat fields. Site identifiers were represented as chr:start0:end1, with the edited nucleotide corresponding to the 1-based end1 coordinate. When multiple transcript-feature annotations were available, one mutually exclusive region label was assigned using the following priority: stop codon, start codon, 3’UTR, 5’UTR, exon, intron, and intergenic. Sites without transcript-feature annotation were classified as intergenic. Alu overlap was defined by a REDIportal type annotation of ALU or a repeat-family annotation containing Alu.

### **Single-site *cis*-edQTL mapping**

For *cis*-edQTL mapping, editing matrices were processed separately by compartment. Sites with >40% missingness or standard deviation <0.005 were removed, remaining missing values were imputed using the site mean, and sites were inverse-normal transformed across samples, following the GTEx edQTL pipeline^S8^.

To infer hidden confounders, PEER factors^S9^ were estimated separately by compartment from the normalized editing matrices. PEER was run with add-mean enabled and fixed covariates age, sex, genotype PCs 1-5, histologic diagnosis and RNA-seq batch. Six hidden PEER factors were included in the final FastQTL covariate files together with fixed covariates, yielding 16 covariates for GLOM and 17 for TUBE.

edQTL mapping was performed using FastQTL^S10^. Variants with MAF ≥ 0.05 were tested within ±100 kb of each editing phenotype. Adaptive permutations used 1,000-10,000 permutations. Site-level q-values were computed from beta-approximated permutation p-values using λ = 0.85, and significant edQTL sites were defined at q < 0.05. Significant variant-site pairs were identified by deriving a global beta-space threshold from the permutation results, converting it to site-specific nominal thresholds, and retaining nominal associations below each site’s threshold.

### **Lead variant proximity**

For each significant single-site edQTL, the lead variant, defined as the variant with the minimum nominal p-value for a given editing phenotype, was obtained from the FastQTL permutation output, and its distance to the editing site was calculated as the absolute genomic distance between the variant and site coordinates. To test whether lead variants were closer to their associated sites than expected, we generated a site-matched null distribution separately in GLOM and TUBE. In each of 10,000 iterations, one tested *cis*-variant was sampled for each significant editing site from that site’s nominal FastQTL variant set, and the mean variant-to-site distance was calculated.

### **Cross-compartment concordance**

Cross-compartment effect concordance was assessed for sites tested in both GLOM and TUBE and significant in at least one compartment. One index variant was selected per site: the lead variant from the significant compartment, or, for sites significant in both compartments, the lead variant from the compartment with the smaller beta-approximated permutation p-value. Nominal FastQTL slopes for the same site-variant pair were extracted from both compartments, and Pearson correlation was used to assess concordance.

### **GWAS and eQTL colocalization**

Colocalization analyses were performed using coloc.abf from the coloc R package with default prior probabilities^S11^. A colocalization event was defined as posterior probability PP.H4 ≥ 0.8. Single-site edQTL-GWAS analyses compared nominal edQTL and GWAS association statistics within the ±100-kb *cis* window centered on each editing site and required at least 25 shared variants after harmonization. cledQTL-GWAS analyses used a ±100-kb window centered on the cluster midpoint, also requiring at least 25 shared variants. GWAS included creatinine-based eGFR^S12^, cystatin C-based eGFR^S13^, blood urea nitrogen^S13^, urinary albumin-to-creatinine ratio^S14^, chronic kidney disease^S15^, IgA nephropathy^S16^, steroid-sensitive nephrotic syndrome^S17^, systolic blood pressure^S18^, and diastolic blood pressure traits^S18^.

Variants were matched by hg38 genomic position and allele. When effect estimates and standard errors were available, GWAS effects were harmonized to the edQTL or cledQTL effect allele. Reverse allele matches were retained after reversing the GWAS effect direction. Ambiguous A/T and C/G variants were excluded. Tests that did not meet the required minimum number of shared variants after harmonization were removed.

Matched-compartment eQTL colocalization used NEPTUNE GLOM eQTL summary statistics for GLOM editing phenotypes and TUBE eQTL summary statistics for TUBE editing phenotypes. For each site-gene or cluster-gene pair, molecular QTL and eQTL variants were harmonized by genomic position and allele. Reverse allele matches were sign-flipped, ambiguous A/T and C/G variants were excluded, and tests with fewer than 50 shared variants were removed.

cledQTL colocalization results were compared with the corresponding single-site edQTL colocalization results. A cledQTL signal was considered to overlap single-site evidence when at least one member editing site in the same cluster showed colocalization with the same GWAS trait or eQTL gene. Signals without corresponding member-site colocalization were classified as cledQTL-only findings.

### **Clinical associations**

Clinical association analyses were performed separately within each tissue compartment and separately for significant single-site edQTLs and significant cledQTL clusters. For single-site edQTLs, linear regression models tested observed editing fraction for association with eGFR at biopsy and log-transformed UPCR. For cledQTLs, analogous linear regression models tested observed cluster mean editing for association with the same continuous outcomes. All linear models adjusted for age, sex, diagnosis, and genotype PCs 1-5. Benjamini-Hochberg FDR correction was applied within tissue and outcome, separately for the single-site edQTL and cledQTL clinical analysis families.

### **CIBERSORTx-derived cell-fraction sensitivity analysis**

Cell-fraction sensitivity analyses used CIBERSORTx^S19^ relative mode with KPMP^S20^-derived broad-cell reference signatures and NEPTUNE compartment-specific bulk RNA-seq mixtures. GLOM and TUBE samples were processed separately using compartment-specific reference signatures. CIBERSORTx was run with quantile normalization disabled, no permutations, and B-mode batch correction. For TUBE sensitivity models, CIBERSORTx selected fractions were proximal tubule, principal cell, thick ascending limb, and immune cell fractions. These cell types were chosen to capture major variable tubular epithelial components and inferred immune-cell abundance.

### **Locuscomparer plots**

Locus comparison plots were generated in R using locuscomparer^S21^. For each comparison, plotted variants were restricted to harmonized variants present in both association datasets within ±100 kb of the QTL locus. Linkage disequilibrium (r^2^) between each plotted variant and a plot-specific reference variant was calculated locally using the function calcld from the gcanvas R package and the corresponding QC-filtered, compartment-specific NEPTUNE PLINK genotype panel^S22^. The calculated LD values were supplied directly to locuscomparer::make_combined_plot().

### **Protein-altering RNA editing annotation**

Protein-altering candidates were drawn from a broader editing-call set before applying the ≥5% sample-presence filter used for the primary FastQTL matrices, to retain rare coding events. Sites were annotated for transcript consequence using VEP (version 115.1)^S23^. Protein-altering consequences included missense variant, stop gained, stop lost, start lost, frameshift variant, protein-altering variant, in-frame insertion or deletion, splice acceptor variant, and splice donor variant.

A candidate was classified as predicted damaging if at least one transcript annotation for that same tissue/site/gene supported SIFT damaging/deleterious, PolyPhen probably/possibly damaging, AlphaMissense likely pathogenic, CADD PHRED ≥20, REVEL ≥0.5, or MPC ≥2 ^S24–S30^. High-confidence predicted damaging candidates required non-conflicting ClinVar pathogenic or likely pathogenic annotation, AlphaMissense likely pathogenic evidence, CADD PHRED ≥25, or REVEL ≥0.75.

### **ClinGen glomerulopathy gene-list prioritization**

Protein-altering candidate events were prioritized using the 157-gene ClinGen Glomerulopathy Gene Curation Expert Panel (GCEP) list^S31^ to identify RNA-editing events in genes with established or proposed glomerulopathy relevance. Candidate events were retained only when the VEP-predicted protein consequence occurred in the same ClinGen-listed gene. GenCC gene-disease metadata^S32^ were added where available for annotation but were not used as an inclusion criterion.

### **Statistical testing**

Statistical analyses were performed in R v4.4.3. Figures were generated using ggplot2 v4.0.0 unless otherwise specified. Two-group comparisons used Wilcoxon rank-sum tests, and multi-group comparisons used Kruskal-Wallis tests. All *P*-values were two-sided.

**SUPPLEMENTARY RESULTS**

### **Protein altering editing events**

Although most human A-to-I editing occurs in noncoding regions, coding edits can alter codons and thereby alter protein sequence or function. Only including editing sites that are in ≥5% of participants is a good strategy for increasing power in QTL mapping, but this filter can exclude rare, deleterious, protein-altering editing events that may still be biologically relevant. To capture these events, we included a set of more rarely observed editing events (218,515 in GLOM, 164,723 in TUBE), with a focus on sites with predicted protein-coding consequences.

This broader candidate set contained 4,690 GLOM and 4,451 TUBE protein-altering editing sites, most of which were annotated as missense events (4,174 GLOM and 4,062 TUBE). Using computational consequence and pathogenicity annotations (**Supplementary Methods**), 2,055 GLOM and 2,130 TUBE sites carried predicted damaging evidence, including 363 and 252 sites, respectively, that met the high-confidence predicted damaging definition.

We next focused on the 157 genes in the ClinGen Glomerulopathy GCEP list^S31^. This yielded 39 observed protein-altering candidates across GLOM and TUBE, 17 of which carried predicted damaging evidence. Four compartment-specific candidates, corresponding to three unique site-gene events, met the high-confidence computational prioritization threshold: Synaptopodin 2 (*SYNPO2)* p.Ser378Gly in GLOM, Phosphoglucomutase 3 (*PGM3)* p.Leu484Pro in GLOM, and Nei-like DNA glycosylase 1 (*NEIL1)* p.Tyr244Cys in both compartments. Several additional candidates had more direct glomerulopathy disease context despite not meeting the same high-confidence computational threshold. These included Tensin 2 (*TNS2;* p.Tyr673Cys), Fibronectin 1 (*FN1;* p.Ser1924Gly), and Crumbs Cell Polarity Complex Component 2 (*CRB2;* p.Thr430Ala), genes implicated in nephrotic syndrome, fibronectin glomerulopathy, and steroid-resistant nephrotic syndrome or FSGS-related phenotypes, respectively. **Supplementary Table 7** summarizes the predicted damaging coding editing candidates together with carrier frequency, editing level, transcript consequence, and disease-context annotations.

These candidates may be useful for targeted follow-up, particularly where editing creates predicted damaging amino acid substitutions in genes with kidney-disease relevance. However, predicted protein consequence alone is not sufficient to infer pathogenicity. Most rare candidates were observed at modest edited-transcript fractions and often with limited edited-read support per carrier, and their interpretation depends on gene-level disease mechanism and mode of inheritance. These results therefore nominate candidate RNA-editing consequences rather than clinical pathogenicity classifications; validation would be required to confirm protein consequence, and phenotypic relevance.

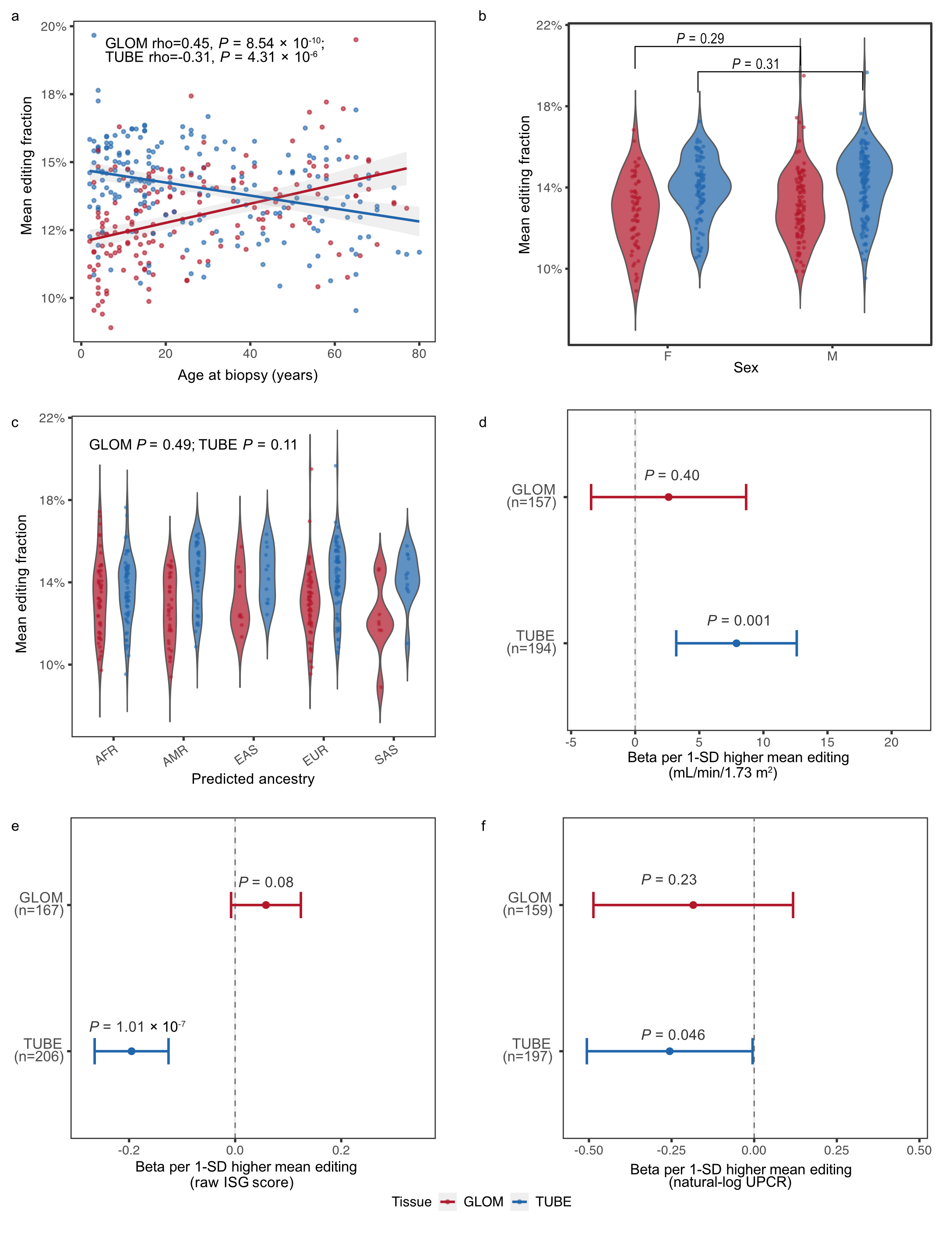

**Supplementary Figure S1. Demographic, clinical, and inflammatory correlates of sample-level mean RNA editing.**

a. Age at biopsy versus per-sample mean editing fraction in GLOM and TUBE. Points represent samples, lines show compartment-specific linear fits, and labels report Spearman correlation coefficients and two-sided *P*-values.

b. Per-sample mean editing fraction stratified by sex and compartment. Violin plots show the distribution of sample-level mean editing, and overlaid points represent individual samples. *P*-values compare female and male samples separately within each compartment using two-sided Wilcoxon rank-sum tests.

c. Per-sample mean editing fraction stratified by genetically inferred ancestry. Violin plots show the distribution of sample-level mean editing, and overlaid points represent individual samples. *P*-values are from Kruskal-Wallis tests across ancestry groups within each compartment. Ancestry abbreviations: AFR, African; AMR, Admixed American; EAS, East Asian; EUR, European; SAS, South Asian.

d.-f. Associations of per-sample mean editing with eGFR at biopsy, ISG-score and natural-log-transformed UPCR at biopsy, respectively. Mean editing was standardized separately within each compartment-specific complete-case analytic sample, and effect estimates are reported per 1-SD higher mean editing. Outcomes remained on their original scales. Models were adjusted for age, sex, and genotype PCs and diagnosis. Points show beta estimates, horizontal bars show 95% confidence intervals, dashed vertical lines indicate the null, and labels report analytic sample sizes and *P*-values for the mean-editing term.

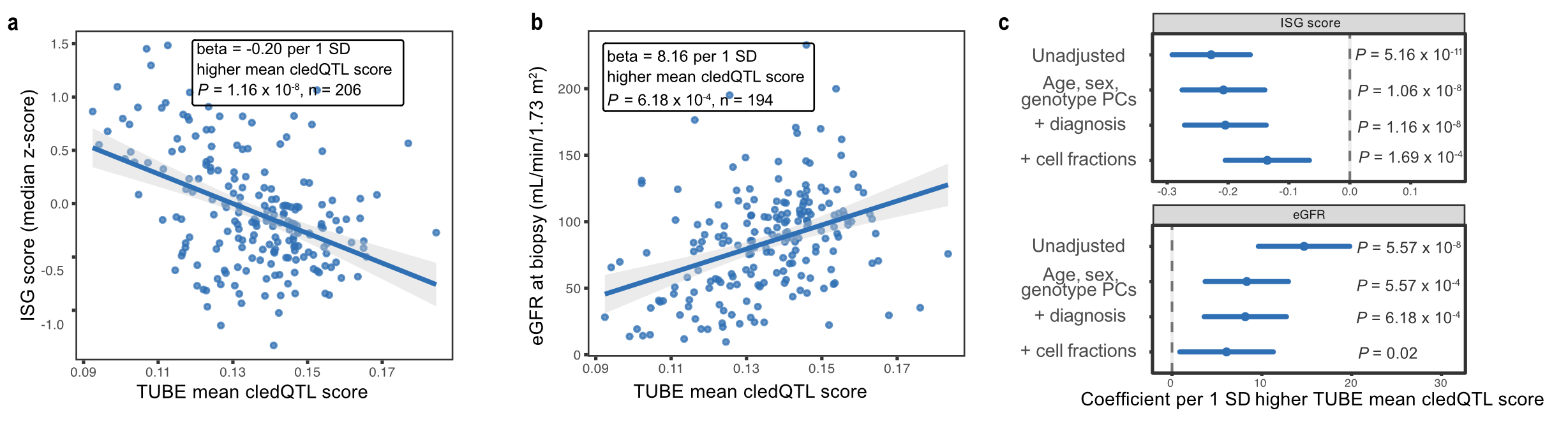

**Supplementary Figure S2. Genetically regulated cluster editing is associated with ISG activity and eGFR.**

a. Association between TUBE mean cledQTL score and interferon-stimulated gene (ISG) activity. The line shows the fitted linear regression with 95% confidence interval. The regression model adjusted for age, sex, genotype PCs, and histologic diagnosis. The coefficient is reported per 1-SD higher TUBE mean cledQTL score.

b. Association between TUBE mean cledQTL score and eGFR at biopsy. The line shows the fitted linear regression with 95% confidence interval. The regression model adjusted for age, sex, genotype PCs, and histologic diagnosis. The displayed coefficient is reported per 1-SD higher TUBE mean cledQTL score.

c. Sensitivity of TUBE mean cledQTL score associations to sequential covariate adjustment. Points show regression coefficients for the association of TUBE mean cledQTL score with ISG-score and eGFR at biopsy; horizontal bars show 95% confidence intervals. Coefficients are reported per 1-SD higher TUBE mean cledQTL score. For ISG models, coefficients are in median ISG z-score units; for eGFR models, coefficients are in mL/min/1.73 m². Rows show unadjusted models and models sequentially adjusted for age, sex, genotype PCs, histologic diagnosis, and CIBERSORTx-derived TUBE cell-fraction estimates for proximal tubule, principal cell, thick ascending limb, and immune fractions. Nominal, two-sided *P*-values from the corresponding linear regression models are reported. The dashed vertical line indicates the null effect.

**Supplementary Table S1. RNA editing matrix construction and completeness summary.**
Summary of RNA editing site filtering, final matrix dimensions, and sample-level completeness for the GLOM and TUBE RNA editing matrices. Editing summary statistics were calculated on the filtered matrix.

**Supplementary Table S2. Genomic annotation of final RNA editing sites.**
Genomic region and Alu repeat annotation of final filtered RNA editing sites in the NEPTUNE FSGS/MCD GLOM and TUBE matrices.

**Supplementary Table S3. Significant cledQTL clusters and variant-cluster associations.**
All significant cluster-level editing QTLs by kidney compartment at FDR q < 0.05. Each row represents a significant cledQTL cluster and reports the lead variant, association statistics, cledQTL effect estimate, MAF, cluster coordinates, number of member sites, cluster span, dominant annotation class, Alu overlap, member site IDs, and overlap with significant single-site edQTL evidence.

**Supplementary Table S4. Clinical associations and colocalization evidence for significant cledQTL clusters.**
Colocalization and clinical association results for all significant cledQTL clusters. Each row represents one cluster and reports the lead cledQTL variant; the number, traits, and top results of GWAS and matched-compartment kidney eQTL colocalizations; and associations with eGFR at biopsy and log-transformed UPCR. Clinical results are provided for the primary covariate-adjusted models and models additionally adjusted for global mean RNA editing, denoted as globally adjusted. Colocalization was defined as PP.H4 ≥ 0.8.

**Supplementary Table S5. Integrated edQTL colocalization and clinical association evidence for significant edQTLs.**
Integrated summary of all significant single-site edQTLs with GWAS colocalization, eQTL colocalization, and/or NEPTUNE clinical association evidence. For each tissue-site-gene entry, the table reports GWAS trait and PP.H4 where applicable, eQTL gene and PP.H4 where applicable, and clinical association results for eGFR and log-transformed UPCR. Clinical results are shown for the primary covariate-adjusted models and for models additionally adjusted for compartment-level global mean RNA editing, denoted as globally adjusted.

**Supplementary Table S6. GWAS datasets used for colocalization.**
GWAS datasets used for cledQTL and edQTL colocalization analyses. The table reports phenotype, publication, year, sample size, ancestry, and download URL.

**Supplementary Table S7. Protein-altering RNA-editing candidates in ClinGen glomerulopathy genes.**
Protein-altering RNA-editing candidates in genes from the ClinGen Glomerulopathy Gene Curation Expert Panel list that were supported by at least one damagingness annotation. Each row represents one tissue-site-gene candidate and reports the edited site, transcript consequence, protein change, predicted damaging evidence, SIFT, PolyPhen, CADD, REVEL, AlphaMissense annotations, tested and edited participant counts, carrier prevalence, carrier editing fractions, GenCC disease-context metadata, and diagnosis counts.

**SUPPLEMENTARY TEXT**

**Members of the Nephrotic Syndrome Study Network (NEPTUNE)**

**NEPTUNE Collaborating Sites**

*Atrium Health Levine Children’s Hospital, Charlotte, SC*: Susan Massengill^*^, Layla Lo^#^

*Cleveland Clinic, Cleveland, OH*: Katherine Dell^*^, John O’Toole^*^, John Sedor^**^, Victoria Grange^#^

*Children’s Hospital, Denver, CO:* Bradley Dixon^*^, Nathan Rogers^#^

*Children’s Hospital, Los Angeles, CA*: Rachel Lestz^*^, Natalie Esquivias^#^

*Children’s Mercy Hospital, Kansas City, MO*: Tarak Srivastava^*^, Kelsey Markus^#^

*Cohen Children’s Hospital, New Hyde Park, NY*: Christine Sethna^*^, Suzanne Vento^#^

*Columbia University, New York, NY:* Pietro Canetta^*^

*Duke University Medical Center, Durham, NC:* Opeyemi Olabisi^*^, Rasheed Gbadegesin^**^, Kimberly Cicio^#^

*Emory University, Atlanta, GA:* Laurence Greenbaum^*^, Chia-shi Wang^*^, Chris Fan^#^

*The Lundquist Institute, Torrance, CA:* Sharon Adler^*^, Janine LaPage^#^

*John H Stroger Cook County Hospital, Chicago, IL:* Amatur Amarah^*^

*Johns Hopkins Medicine, Baltimore, MD:* Meredith Atkinson^*^, Ryan Hutson^#^

*Mayo Clinic, Rochester, MN:* John Lieske, Marie Hogan, Fernando Fervenza

*Medical University of South Carolina, Charleston, SC:* David Selewski^*^, Cheryl Alston^#^

*Montefiore Medical Center, Bronx, NY:* Kim Reidy^*^, Michael Ross^*^, Frederick Kaskel^**^, Patricia Flynn^#^

*New York University Medical Center, New York, NY:* Laura Malaga-Dieguez^*^, Olga Zhdanova^**^, Laura Jane Pehrson^#^, Melanie Miranda^#^

*The Ohio State University College of Medicine, Columbus, OH*: Salem Almaani^*^, Laci Roberts^#^

*Riley Children’s Hospital of Indiana University, Indianapolis, IN:* Myda Khalid^*^, Veronica Servin^#^

*Stanford University, Stanford, CA:* Richard Lafayette^*^, Elizabeth Chen^#^

*Temple University, Philadelphia, PA:* Iris Lee^**^

*Texas Children’s Hospital at Baylor College of Medicine, Houston, TX*: Shweta Shah^*^, Thinh Phan^#^

*University Health Network Toronto:* Heather Reich^*^, Michelle Hladunewich^**^, Paul Ling^#^, Martin Romano^#^

*University of California at San Diego, San Diego, CA:* Ambarish Athavale^*^, Caitlin Carter^*^, Kristin Zeeb^#^

*University of California at San Francisco, San Francisco, CA*: Paul Brakeman^*^, Daniel Schrader

*University of Colorado Anschutz Medical Campus, Aurora, CO*: James Dylewski^*^ Nathan Rogers^#^

*University of Kansas Medical Center, Kansas City, KS*: Ellen McCarthy^*^, Catherine Creed^#^

*University of Miami, Miami, FL:* Alessia Fornoni^*^, Miguel Bandes^#^

*University of Michigan, Ann Arbor, MI:* Matthias Kretzler^*^, Laura Mariani^*^, Zubin Modi^*^, Amanda Williams^#^, Roxy Ni^#^

*University of Minnesota, Minneapolis, MN:* Patrick Nachman^*^, Michelle Rheault^*^, Ariel Langenberger^#^, Brady Wallner^#^

*University of North Carolina, Chapel Hill, NC:* Vimal Derebail^*^, Keisha Gibson^*^, Anne Froment^#^, Sharia Warren^#^

*University of Pennsylvania, Philadelphia, PA:* Lawrence Holzman^*^, Kevin Meyers^**^, Krishna Kallem^#^, Arielle Swenson^#^

*University of Texas San Antonio, San Antonio, TX*: Samin Sharma^**^

*University of Texas Southwestern, Dallas, TX:* Elizabeth Roehm^*^, Kamalanathan Sambandam^**^, Elizabeth Brown^**^

*University of Washington, Seattle, WA:* Ashley Jefferson^*^, Sangeeta Hingorani^**^, Katherine Tuttle^**§^, Linda Manahan ^#^, Emily Pao^#^, Kelli Kuykendall^§^

*Wake Forest University Baptist Health, Winston-Salem, NC:* Jen Jar Lin^**^

*Washington University in St. Louis, St. Louis, MO*: Brian Stotter^*^, Joseph Dumayas^#^

**Data Analysis and Coordinating Center:** *University of Michigan:* Matthias Kretzler^*^, Brenda Gillespie^**^, Laura Mariani^**^, Zubin Modi^**^, Eloise Salmon^**^, Howard Trachtman^**^, Hailey Desmond, Sean Eddy, Damian Fermin, Wenjun Ju, Maria Larkina, Chrysta Lienczewski, Rebecca Scherr, Jonathan Troost, Amanda Williams, Yan Zhai; *Cleveland Clinic:* Crystal Gadegbeku^**^, John Sedor^**^, *Duke University:* Laura Barisoni^**^; *Harvard University:* Matthew G Sampson^**^; *Northwestern University:* Abigail Smith^**^; *University of Pennsylvania:* Lawrence Holzman^**^, Jarcy Zee^**^

**Digital Pathology Committee:** Carmen Avila-Casado *(University Health Network)*, Serena Bagnasco *(Johns Hopkins University)*, Lihong Bu *(Mayo Clinic)*, Shelley Caltharp *(Emory University)*, Clarissa Cassol *(Arkana)*, Dawit Demeke *(University of Michigan)*, Brenda Gillespie *(University of Michigan)*, Jared Hassler *(Temple University)*, Leal Herlitz *(Cleveland Clinic)*, Stephen Hewitt *(National Cancer Institute)*, Jeff Hodgin *(University of Michigan)*, Danni Holanda *(Arkana)*, Neeraja Kambham *(Stanford University)*, Kevin Lemley, Laura Mariani *(University of Michigan)*, Nidia Messias *(Washington University)*, Alexei Mikhailov *(Wake Forest)*, Vanessa Moreno *(University of North Carolina)*, Behzad Najafian *(University of Washington)*, Matthew Palmer *(University of Pennsylvania)*, Avi Rosenberg *(Johns Hopkins University)*, Virginie Royal *(University of Montreal)*, Miroslav Sekulik *(Columbia University)*, Barry Stokes *(Columbia University)*, David Thomas *(Duke University)*, Ming Wu *(University of New York)*, Michifumi Yamashita *(Cedar Sinai)*, Hong Yin *(Emory University)*, Jarcy Zee *(University of Pennsylvania)*, Yiqin Zuo *(University of Miami)*. Co-Chairs: Laura Barisoni *(Duke University)*, Cynthia Nast *(Cedar Sinai)*.
